# Revealing Hidden Myocardial Infarction Signatures from Brief Single-Lead Electrocardiograms: A Novel Framework for Smart Wearable Applications

**DOI:** 10.64898/2026.07.08.26357521

**Authors:** Rashid Alavi, Jiajun Li, Ray V. Matthews, Niema M. Pahlevan, Robert A. Kloner, Morteza Gharib

**Author notes:** **Corresponding Authors:** - Morteza Gharib, PhD, - Robert A. Kloner, MD, PhD.

## Abstract

The electrocardiogram (ECG) contains rich nonlinear and non-stationary dynamic information that is only partly captured by conventional ECG interpretation and beat-to-beat metrics, and is increasingly analyzed using black-box artificial intelligence models that often lack interpretability. Here, we introduce the ECG time–frequency “eyeball”, an interpretable framework that transforms a brief single-lead ECG recording into a geometric signature and a set of low-dimensional rotational and geometrical features using empirical mode decomposition and Hilbert-based analytic signal mapping. In 30-second Lead I–equivalent recordings from 170 healthy subjects and 80 patients with acute myocardial infarction (AMI), the proposed “eyeball” metrics significantly differentiated groups, with AMI associated with higher rotational frequency metrics, lower envelope metrics, and displaced centroid location. Representative examples revealed a coherent morphologic spectrum from normal patterns to geometries consistent with myocardial ischemia, injury, and infarction. The representation remained stable across recording windows from 30 seconds to 5 minutes, and individual “eyeball” features achieved areas under the receiver operating characteristic curve (AUCs) of up to 0.78 for AMI detection. These findings suggest that the ECG time–frequency “eyeball” condenses clinically relevant nonlinear ECG dynamics into an interpretable representation that may reveal hidden AMI signatures, complement conventional ECG interpretation, and provide a foundation for accessible single-lead cardiovascular screening using future smart wearables.

## Introduction

Surface electrocardiography (ECG) remains one of the most widely deployed diagnostic tests in cardiovascular medicine and continues to play a crucial role in the early detection and evaluation of acute coronary syndromes, particularly acute myocardial infarction (AMI). Yet conventional ECG interpretation still relies predominantly on visible time-domain landmarks (e.g., ST-segment deviation, T-wave inversion, QRS distortion, and interval measurements) and on their integration with symptoms and biomarkers. Although these features are clinically indispensable, manifestations of myocardial ischemia and infarction are often subtle, distributed throughout the signal, and dynamically evolving, suggesting that important diagnostic information may remain hidden from conventional ECG interpretation^1–5^.

Recent advances in artificial intelligence (AI) have reinforced this view. Deep-learning analysis of raw ECGs can identify reduced ejection fraction, hypertrophic cardiomyopathy, structural heart disease and acute coronary syndromes, often from waveforms whose full diagnostic content is not captured by routine interpretation^6–13^. These studies suggest that the ECG contains a richer latent description of cardiac physiology and pathology than is captured by conventional rule-based reading. However, most AI-ECG systems still function primarily as prediction engines, offering limited intuitive visibility into the underlying signal organization that drives their outputs.

This interpretability gap becomes even more important as ECG acquisition moves beyond the clinic. Smartwatches, handheld electrodes, and other smart wearable platforms can now acquire brief single-lead ECGs at scale, and recent studies have shown their utility for arrhythmia screening as well as for the identification of structural cardiac phenotypes^13–19^. Yet such recordings are short and reduced in dimensionality, and are often acquired under variable conditions. What is needed are methods that preserve clinically meaningful information in brief single-lead recordings while remaining interpretable enough for clinical inspection and scalable enough for digital or wearable deployment.

Time–frequency analysis offers a principled route to such a representation because cardiac electrical activity is intrinsically nonlinear and non-stationary. Empirical mode decomposition and Hilbert spectral analysis can adaptively decompose complex biosignals into intrinsic oscillatory components and quantify their instantaneous amplitude and frequency without imposing stationarity assumptions^20–22^. In practical terms, this allows dynamic electrical fluctuations embedded within the ECG to be tracked continuously over time, rather than being reduced to static interval or amplitude measurements from a few fiducial points. Such approaches may therefore capture subtle electrophysiological alterations that are not readily visible using conventional waveform inspection alone. More broadly, time–frequency methods have been applied to ECG denoising, feature extraction and automated classification, and geometric or phase-space approaches have suggested that myocardial ischemia perturbs global ECG morphology in ways not fully captured by conventional waveform rules^21,23^. Nonetheless, most prior approaches either remain mathematically abstract or reduce the signal to features that are not directly visualizable, limiting bedside interpretability and translational appeal.

Here, we introduce the ECG time–frequency “eyeball”, an interpretable framework that transforms a brief single-lead ECG recording into a stable geometric representation and a set of low-dimensional rotational and geometrical features. By decomposing the recording into intrinsic mode functions, constructing their analytic signals, and superposing the corresponding complex-plane trajectories, the framework reveals hidden nonlinear and dynamically evolving ECG information without requiring conventional waveform delineation or reliance on predefined ECG waveform abnormalities. Using brief lead-I–equivalent recordings from healthy individuals and patients with acute myocardial infarction (AMI), we show that the framework reveals hidden AMI signatures, organizes representative morphologies across a spectrum of myocardial conditions, and remains robust for recordings as short as 30 seconds. In doing so, it offers a potential bridge between classical ECG interpretation and modern data-driven cardiovascular diagnostics, paving the way for accessible cardiovascular screening using smart wearable devices.

## Results

### Study Population

A total of 250 subjects were included in the analysis, comprising 170 healthy individuals and 80 patients with acute myocardial infarction (AMI) (Table 1). The AMI cohort was older than the healthy cohort (57.0 ± 14.1 vs. 39.0 ± 14.2 years) and had a higher proportion of males (77.5% vs. 50%). Other baseline characteristics, including body mass index, heart rate, and blood pressure, were generally comparable between groups.

**Table 1.** Characteristics of the study population.

| Characteristic | Healthy (N=170) | AMI (N=80) |
| --- | --- | --- |
| **Sex, n(%)** |  |  |
| Male | 85 (50%) | 62 (77.5%) |
| Female | 85 (50%) | 18 (22.5%) |
| **Age (years)** | 18-80 ($39.0 \pm 14.2$) | 27-90 ($57.0 \pm 14.1$) |
| **BMI ($\text{kg}/\text{m}^2$)\*** | 17.28-49.61 ($24.4 \pm 4.5$) | 18.36-43.44 ($26.7 \pm 3.7$) |
| **Height (cm)** | 149-196 ($170.2 \pm 10.2$) | 150-188 ($169.5 \pm 8.0$) |
| **Weight (kg)** | 45-127 ($70.9 \pm 15.3$) | 47-114 ($76.9 \pm 12.9$) |
| **Heart Rate (bpm)** | 41.7-107.3 ($67.5 \pm 14.4$) | 48.4-106.7 ($69.6 \pm 12.9$) |
| **Blood Pressure (mmHg)\*\*** |  |  |
| Systolic | 90-180 ($117 \pm 13$) | 90-180 ($124 \pm 16$) |
| Diastolic | 60-100 ($76 \pm 8$) | 55-120 ($78 \pm 10$) |

Data are presented as minimum–maximum (mean ± standard deviation) unless otherwise noted.
\*Height and weight measurements were unavailable for one individual in the Healthy database.
\*\*Blood pressure statistics were calculated excluding 18 individuals in the Healthy database without measurements.

### ECG Time–Frequency “Eyeball” Representation

The workflow for constructing the ECG time–frequency “eyeball” from 30-second single-lead recordings is illustrated in Figure 1. Decomposition of ECG signals into intrinsic mode functions (IMFs), followed by Hilbert-based analytic signal construction and rotational mapping, yields trajectories in the complex plane. Superposition of these trajectories from the first four IMFs produces a consistent “eyeball”-like geometry across subjects, enabling extraction of rotational and geometrical features.

**Figure 1.**
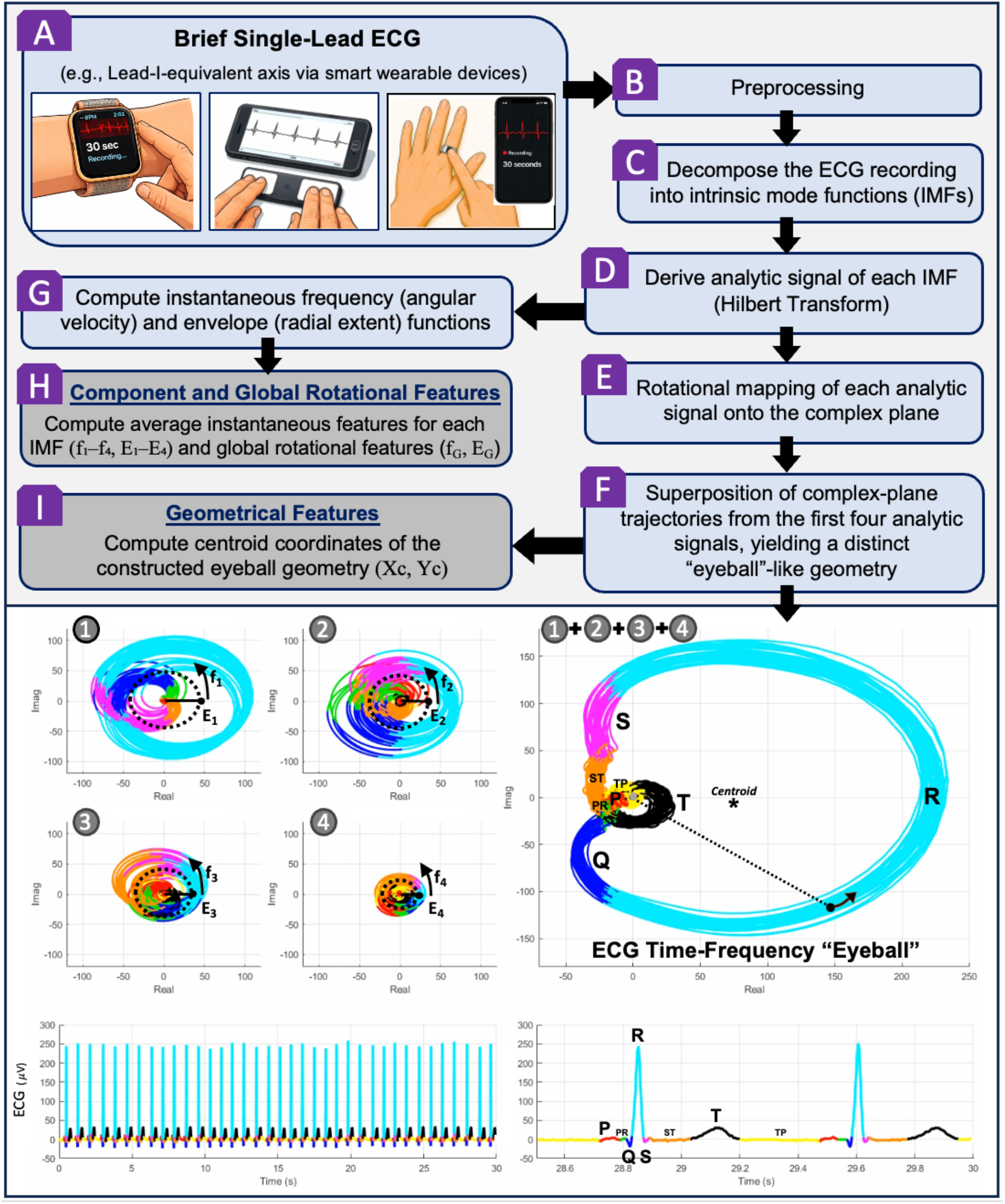
Methodology overview and workflow for constructing the ECG time–frequency “eyeball” from a brief single-lead recording. **Top Panel: (A)** Acquisition of a brief single-lead ECG (e.g., Lead-I–equivalent axis via smart wearable devices or handheld devices). **(B)** ECG signal preprocessing, including standardized resampling and filtering. **(C–D)** Decomposition of the ECG recording into its intrinsic mode functions (IMFs) using empirical mode decomposition, followed by derivation of the analytic signal for each IMF (via Hilbert transform). **(E)** Rotational mapping of each analytic signal onto the complex plane. **(F)** Superposition of complex-plane trajectories from the first four analytic signals, yielding a distinct “eyeball”-like geometry (termed the “ECG time–frequency eyeball”). **(G)** Computation of instantaneous rotational features for each IMF trajectory, including instantaneous frequency (i.e., angular velocity) and instantaneous envelope (i.e., radial extent). **(H)** Defining the eyeball’s rotational features including component and global frequencies and envelopes: Computation of average instantaneous features for each IMF (f_1_-f_4_, E_1_-E_4_) and global rotational features (f_G_, E_G_). **(I)** Defining the eyeball’s geometrical features: Quantification of geometric descriptors of the constructed “eyeball”, including centroid coordinates (X_C_, Y_C_). **Bottom Panel:** Representative visualization of the ECG time–frequency “eyeball” for a healthy subject is shown, where the first four intrinsic mode functions (*IMF*_1_(*t*) to *IMF*_4_(*t*)) are rotationally mapped onto the complex plane using their analytic signals (real vs. imaginary components) and the corresponding rotational features (instantaneous frequency and instantaneous envelope). Their progressive superposition forms the “eyeball” geometry, illustrating the emergence of a stable morphological structure (Supplementary Movie 1). Here, *f*_*i*_ and *E*_*i*_ (*i* =1-4) denote the component frequencies and envelopes of the ECG “eyeball”, respectively. Both the accumulated ECG waveforms (full 30-second recording) and the live ECG signal (last two waveforms) are displayed, highlighting the correspondence between ECG components and their “eyeball” representations. ECG components are color-coded as follows: P wave (red), PR segment (green), Q wave (dark blue), R wave (cyan), S wave (purple), ST segment (orange), T wave (black), and TP segment (yellow).

The dynamic formation of the “eyeball” geometry from a 30-second ECG recording is illustrated in a synchronized animated visualization (Supplementary Movie 1), demonstrating the progressive emergence of a stable morphological structure through the superposition of IMF trajectories.

### Distribution of ECG “Eyeball” Metrics in Healthy and AMI Cohorts

Significant differences were observed in ECG “eyeball” metrics between healthy and AMI cohorts (Figure 2, Table 2). Compared to healthy subjects, AMI patients exhibited consistently higher “eyeball” frequencies (f_1_–f_4_, f_G_), marked reductions in “eyeball” envelopes (E_1_–E_4_, E_G_), and substantial shifts in “eyeball’s” centroid location (X_C_, Y_C_). For example, mean f_G_ increased from 217.0 bpm in healthy subjects to 256.4 bpm in AMI (p<0.0001), while E_G_ decreased from 34.68 μV to 24.07 μV (p<0.0001; Table 2). Similarly, the mean centroid coordinates (X_C_, Y_C_) decreased from (28.97, 4.37) in healthy subjects to (10.25, 1.42) in the AMI cohort (p<0.0001 and p<0.01, respectively), indicating a pronounced geometric shift. These differences were statistically significant for all features (Figure 2A, 2B, 2C; Table 2), indicating systematic alterations in both rotational and geometrical properties of the “eyeball” representation in AMI.

**Table 2.** Summary statistics of metrics derived from the ECG time–frequency “eyeball” framework in healthy and acute myocardial infarction (AMI) cohorts.

| Variable | Healthy |  |  | AMI |  |  | p-value |
| --- | --- | --- | --- | --- | --- | --- | --- |
|  | Mean | Median | IQR | Mean | Median | IQR |  |
| $f_1$ (bpm) | 538.6 | 426.9 | [276.6, 662.4] | 834.2 | 674.3 | [409.6, 1050.8] | <0.0001 |
| $f_2$ (bpm) | 289.8 | 250.7 | [210.1, 319.2] | 379.7 | 349.4 | [273.0, 433.6] | <0.0001 |
| $f_3$ (bpm) | 166.2 | 149.9 | [132.5, 192.0] | 191.3 | 182.8 | [160.1, 210.4] | <0.0001 |
| $f_4$ (bpm) | 74.1 | 67.8 | [58.5, 85.9] | 90.4 | 87.9 | [73.9, 101.9] | <0.0001 |
| $f_G$ (bpm) | 217.0 | 195.8 | [164.5, 255.1] | 256.4 | 245.3 | [198.4, 300.7] | <0.0001 |
| $E_1$ ($\mu V$) | 13.02 | 12.06 | [9.22, 15.95] | 8.16 | 7.62 | [5.12, 10.72] | <0.0001 |
| $E_2$ ($\mu V$) | 17.40 | 16.34 | [12.47, 22.08] | 10.57 | 10.23 | [6.41, 13.64] | <0.0001 |
| $E_3$ ($\mu V$) | 15.58 | 14.72 | [11.18, 19.47] | 12.00 | 11.59 | [7.80, 15.11] | <0.0001 |
| $E_4$ ($\mu V$) | 12.71 | 12.13 | [8.34, 16.15] | 9.97 | 8.79 | [5.93, 13.11] | <0.001 |
| $E_G$ ($\mu V$) | 34.68 | 33.00 | [25.14, 44.03] | 24.07 | 22.32 | [16.29, 30.54] | <0.0001 |
| $X_C$ ($\mu V$) | 28.97 | 27.36 | [16.56, 40.39] | 10.25 | 7.81 | [-1.39, 20.78] | <0.0001 |
| $Y_C$ ($\mu V$) | 4.37 | 3.71 | [-1.56, 11.09] | 1.42 | 0.56 | [-4.17, 5.30] | <0.01 |

**Figure 2.**
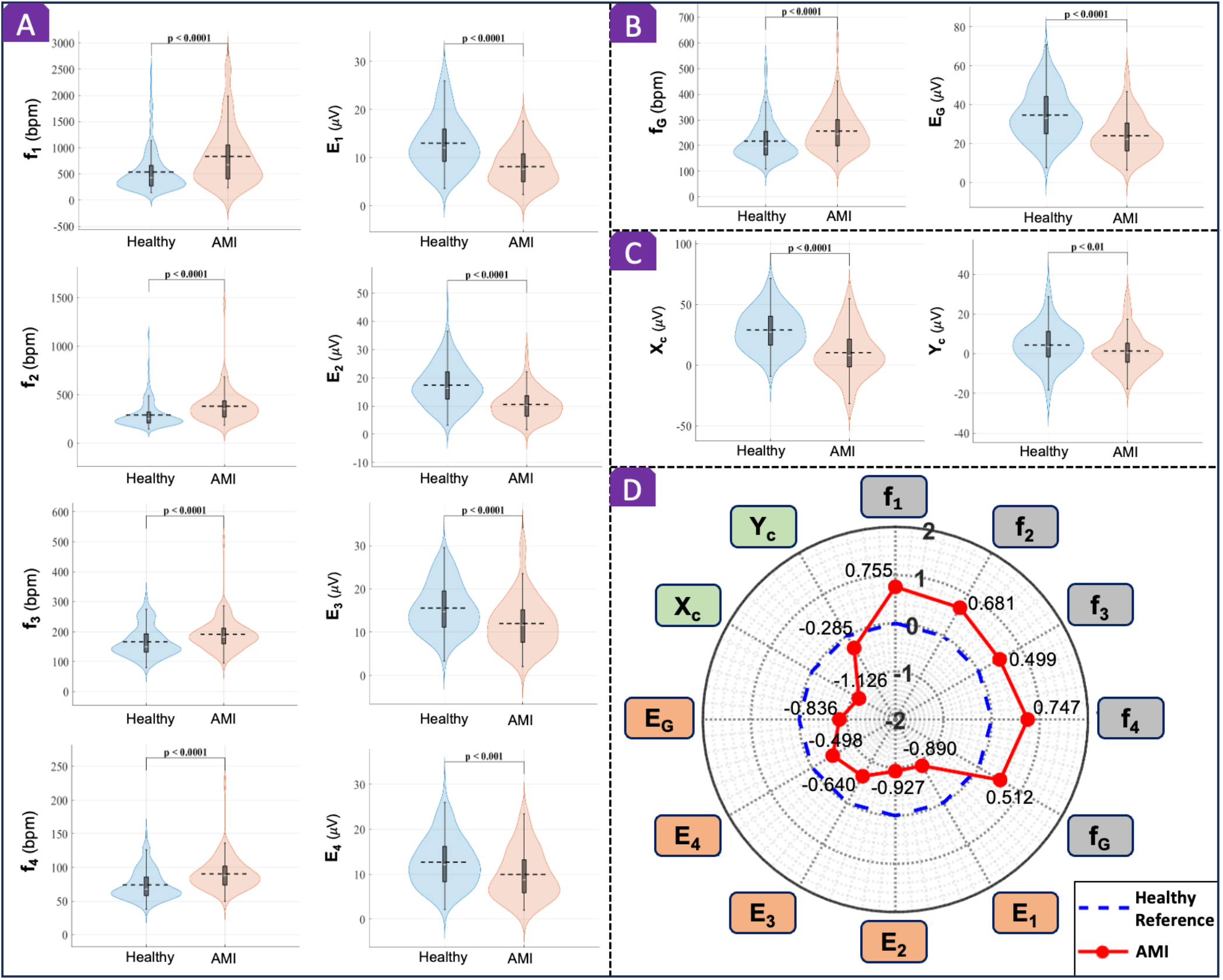
Distribution of ECG time–frequency “eyeball” metrics in healthy versus acute myocardial infarction (AMI) cohorts. **(A, B, C)** Violin plots overlaid with box-and-whisker plots comparing the distribution of proposed metrics computed from 30-second single-lead ECG recordings in healthy subjects and patients with AMI. Dashed lines indicate mean values, while the central white line within each box denotes the median. Box-and-whisker plots also represent the interquartile range (IQR). Differences between healthy and AMI cohorts for each metric were assessed using an unpaired t-test or the Mann–Whitney U test, as appropriate, with corresponding p-values indicated in the figure. **(A)** Component rotational features: *f*_*i*_ and *E*_*i*_ (*i* =1–4) represent the component frequencies and envelopes of the ECG “eyeball”, respectively. **(B)** Global rotational features: *f*_*i*_ and *E*_*i*_ are the global ECG “eyeball” frequency and envelope, respectively. **(C)** Geometrical features: X_G_ and Y_G_ denote the centroid coordinates of the “eyeball” geometry. **(D)** Radar plot representation of proposed “eyeball” metrics illustrating the group-averaged normalized values and deviation of AMI cohort from the healthy reference. Metrics were normalized relative to the healthy cohort using z-score normalization (mean and standard deviation of the healthy group), such that the healthy reference is centered at zero (blue dashed line), and the AMI cohort (red line) reflects deviations from this baseline across all components. This visualization highlights the coordinated increase in ECG “eyeball” frequencies, reduction in ECG “eyeball” envelopes, and shifts in “eyeball’s” centroid location, revealing a distinct multidimensional signature of AMI.

Radar plot visualization of the ECG “eyeball” metrics further highlights a coordinated multidimensional signature of AMI, characterized by a systematic shift toward higher component and global frequencies (f_1_–f_4_, f_G_), lower component and global envelopes (E_1_–E_4_, E_G_), and displacement of the “eyeball” centroid (X_C_, Y_C_) relative to the healthy reference (Figure 2D).

*f*_*i*_ and *E*_*i*_ (*i* = 1-4) denote the component frequencies and envelopes of the ECG “eyeball”, respectively. *f*_*G*_ and *E*_*G*_ are the global ECG “eyeball” frequency and envelope, respectively. (X_C_, Y_C_): Centroid coordinates of the “eyeball” geometry; IQR: interquartile range. P-values represent comparisons between healthy and AMI cohorts using an unpaired t-test or Mann– Whitney U test, as appropriate.

### Association of ECG “Eyeball” Metrics with AMI

Logistic regression analysis demonstrated significant associations between ECG “eyeball” metrics and the odds of AMI (Figure 3A; Table 3). In unadjusted models, all “eyeball” frequencies (f_1_–f_4_, f_G_) were positively associated with AMI (OR_n_ > 1), while the “eyeball” envelopes (E_1_–E_4_, E_G_) and centroid coordinates (X_C_, Y_C_) showed inverse associations (OR_n_ < 1).

**Table 3.** Logistic regression analysis results for models of acute myocardial infarction (AMI)

| Variable | Unadjusted Models |  | Adjusted Models<br>(age and sex) |  | Adjusted Models<br>(age, sex, and HR) |  |
| --- | --- | --- | --- | --- | --- | --- |
|  | OR<sub>n</sub> [95% CI] | AUC | OR<sub>n</sub> [95% CI] | p-value | OR<sub>n</sub> [95% CI] | p-value |
| **f<sub>1</sub>** | 1.872 [1.419, 2.533] | 0.695 | 1.573 [1.120, 2.245] | 0.0086* | 1.462 [1.030, 2.100] | 0.0334* |
| **f<sub>2</sub>** | 1.936 [1.420, 2.720] | 0.724 | 1.490 [1.065, 2.180] | 0.0186* | 1.340 [0.941, 1.993] | 0.1077 |
| **f<sub>3</sub>** | 1.617 [1.228, 2.167] | 0.656 | 1.398 [1.002, 2.003] | 0.0487* | 1.215 [0.833, 1.810] | 0.3162 |
| **f<sub>4</sub>** | 2.095 [1.555, 2.884] | 0.725 | 1.812 [1.275, 2.647] | 0.0007* | 1.689 [1.139, 2.572] | 0.0086* |
| **f<sub>G</sub>** | 1.614 [1.236, 2.140] | 0.665 | 1.465 [1.059, 2.062] | 0.0208* | 1.311 [0.911, 1.902] | 0.1436 |
| **E<sub>1</sub>** | 0.274 [0.176, 0.409] | 0.765 | 0.460 [0.281, 0.723] | 0.0005* | 0.462 [0.280, 0.731] | 0.0007* |
| **E<sub>2</sub>** | 0.258 [0.164, 0.387] | 0.782 | 0.412 [0.245, 0.664] | 0.0002* | 0.428 [0.253, 0.693] | 0.0004* |
| **E<sub>3</sub>** | 0.498 [0.357, 0.677] | 0.683 | 0.688 [0.465, 1.006] | 0.0537 | 0.704 [0.472, 1.038] | 0.0764 |
| **E<sub>4</sub>** | 0.581 [0.424, 0.779] | 0.648 | 0.708 [0.487, 1.013] | 0.0592 | 0.764 [0.519, 1.112] | 0.1608 |
| **E<sub>G</sub>** | 0.345 [0.233, 0.491] | 0.744 | 0.516 [0.327, 0.789] | 0.0019* | 0.535 [0.337, 0.824] | 0.0041* |
| **X<sub>C</sub>** | 0.298 [0.200, 0.426] | 0.772 | 0.515 [0.335, 0.770] | 0.0011* | 0.548 [0.354, 0.826] | 0.0039* |
| **Y<sub>C</sub>** | 0.737 [0.552, 0.970] | 0.606 | 0.712 [0.500, 0.999] | 0.0493* | 0.703 [0.490, 0.995] | 0.0468* |

Values are presented as normalized odds ratios (OR<sub>n</sub>) with 95% confidence intervals (95% CI). OR<sub>n</sub> represents the odds ratio associated with a one-standard deviation increase in each ECG “eyeball” feature (i.e., how do the odds of AMI change, holding covariates constant). Area under the receiver operating characteristic curve (AUC) values correspond to ROC analyses performed using each ECG “eyeball” feature individually (unadjusted models). Adjusted models included age and sex, or age, sex, and heart rate (HR) as covariates. For adjusted models, p-values were obtained using the likelihood-ratio test comparing a reduced model containing only the clinical covariates with a full model that additionally included the ECG “eyeball” feature. The null hypothesis was that addition of the ECG “eyeball” feature did not improve model fit. Statistical significance was defined as $p < 0.05$ (\*). $f_i$ and $E_i$ ( $i = 1-4$ ) denote the component frequencies and envelopes of the ECG “eyeball”, respectively. $f_G$ and $E_G$ are the global ECG “eyeball” frequency and envelope, respectively. ( $X_C$ , $Y_C$ ): Centroid coordinates of the “eyeball” geometry.

**Figure 3.**
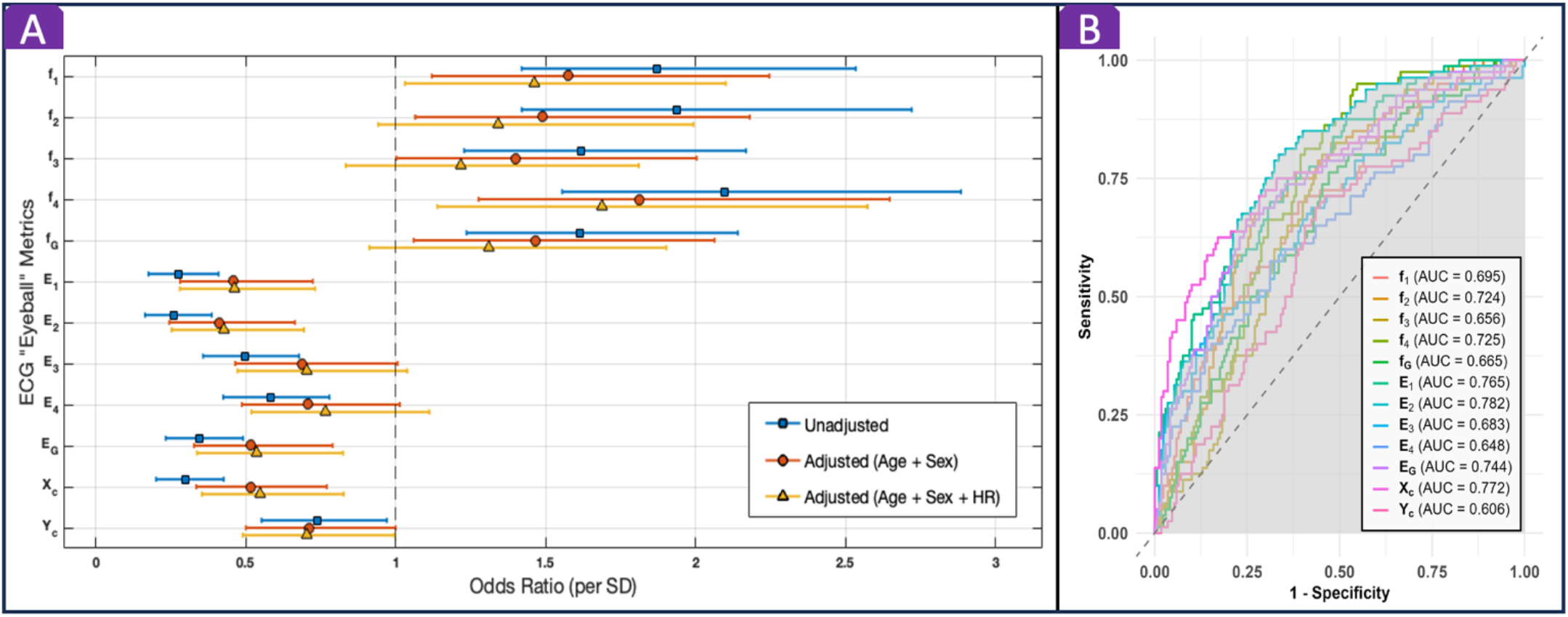
Association and discriminative performance of ECG “eyeball” metrics for acute myocardial infarction (AMI). **(A)** Association of each ECG “eyeball” metric with the odds of AMI. The forest plot presents normalized odds ratios (OR_n_; odds ratio per standard deviation [SD] increase in each metric) derived from logistic regression models. Three logistic regression models are shown for each metric: unadjusted, adjusted for age and sex, and further adjusted for heart rate (HR). Points indicate OR_n_ estimates, and horizontal lines represent 95% confidence intervals. The vertical dashed line denotes OR_n_ = 1 (no association with AMI). **(B)** Receiver operating characteristic (ROC) curves illustrating the discriminative performance of each individual ECG “eyeball” metric for distinguishing patients with AMI from healthy subjects. The area under the curve (AUC) is reported for each metric. *f*_*i*_ and *E*_*i*_ (*i* = 1-4) represent the component frequencies and envelopes of the ECG “eyeball”, respectively. *f*_*G*_ and *E*_*G*_ are the global ECG “eyeball” frequency and envelope, respectively. X_C_ and Y_C_ denote the centroid coordinates of the “eyeball” geometry.

After adjustment for age, sex, and heart rate, the overall pattern of associations was preserved, with “eyeball” frequencies remaining positively associated with AMI, and “eyeball” envelopes and centroid coordinates showing inverse relationships (Figure 3A; Table 3). Features remained independently associated with AMI and provided significant incremental value beyond clinical covariates, including f_1_ (OR_n_ = 1.462, p = 0.0334), f_4_ (OR_n_ = 1.689, p = 0.0086), E_1_ (OR_n_ = 0.462, p = 0.0007), E_2_ (OR_n_ = 0.428, p = 0.0004), E_G_ (OR_n_ = 0.535, p = 0.0041), X_C_ (OR_n_ = 0.548, p = 0.0039), and Y_C_ (OR_n_ = 0.703, p = 0.0468) (Figure 3A; Table 3).

These findings indicate that both rotational and geometrical features of the ECG “eyeball” representation contribute independently to distinguishing AMI from healthy subjects. Adjustments were performed to account for the potential influence of age, sex, and heart rate on ECG-derived features. To further assess age-related effects, associations between proposed “eyeball” metrics and age, including group-dependent differences, were evaluated using multivariable linear regression (Supplementary Table 1).

### Discriminative Performance of ECG “Eyeball” Metrics

The discriminative performance of each ECG “eyeball” metric was evaluated using receiver operating characteristic (ROC) analysis, with the area under the curve (AUC) as the performance metric (Figure 3B; Table 3). Individual ECG “eyeball” metrics demonstrated consistent discriminative ability for identifying AMI, with AUC values ranging from 0.61 to 0.78. Component and global frequency metrics achieved AUCs of approximately 0.66–0.73, component and global envelope metrics demonstrated the highest discriminative performance (AUC ≈ 0.65–0.78), and centroid coordinates yielded AUCs of approximately 0.61–0.77. These findings indicate that individual ECG “eyeball” features capture meaningful discriminatory information for distinguishing patients with AMI from healthy subjects.

### Stability of ECG “Eyeball” Metrics Across Recording Durations

To evaluate the temporal robustness of the proposed framework, all “eyeball” metrics were computed across ECG recordings of varying durations (i.e., 30 seconds, 1 minute, 2 minutes, and 5 minutes). This analysis is particularly relevant for smart wearable applications, where ECG recordings are often brief and acquired intermittently. In both healthy and AMI cohorts, the “eyeball” metrics demonstrated minimal variation across recording durations, reflected by consistently high p-values across all comparisons (all p≥0.93, predominantly p>0.99; see Supplementary Figure 1 and Supplementary Figure 2). These findings support the practical use of brief ECG recordings (30-second) for reliable estimation of the “eyeball” features (both rotational and geometrical features).

Consistent with these quantitative results, representative examples from a healthy subject show that the overall “eyeball” structure remains preserved across increasing durations (30 seconds, 1 minute, 2 minutes, 5 minutes), with only minor variations in trajectory density due to longer signal accumulation (Supplementary Figure 3).

### Morphological Changes of ECG “Eyeball” Geometry Across Myocardial Conditions

Representative examples of ECG “eyeball” geometry across recognizable myocardial conditions are shown in Figure 4. A clear pattern is observed from healthy to acute myocardial infarction (AMI), characterized by a progressive reduction in overall “eyeball” size, alterations in geometric shape and compactness, and increasing irregularity in trajectory structure. The dynamic formation of the representative ECG “eyeballs” shown in Figure 4 is illustrated in Supplementary Movies 2–8, which show the synchronized evolution of ECG waveforms and the corresponding ECG “eyeball” throughout the 30-second recording.

**Figure 4.**
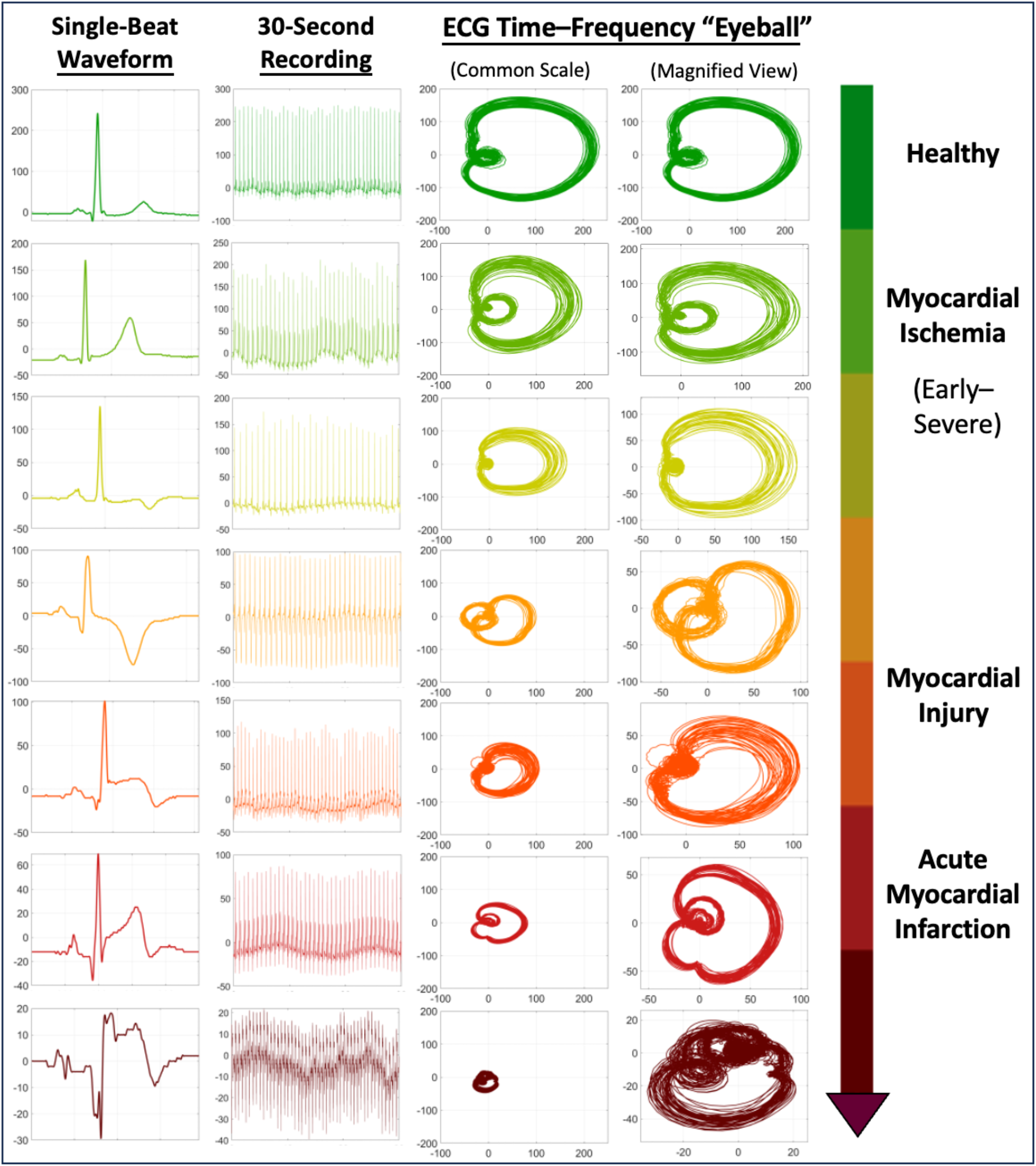
Morphological changes in ECG time–frequency “eyeball” geometry across myocardial conditions. Representative ECG recordings and their corresponding “eyeball” geometry (derived from 30-second recordings) are shown across a spectrum of myocardial conditions, from healthy to acute myocardial infarction (AMI). The “eyeball” geometry is displayed using both a common scale (third column) to enable direct comparison of global size and a magnified view (fourth column) to highlight morphological details. All common-scale “eyeball” plots share identical axis limits, demonstrating a progressive reduction in “eyeball” size from healthy to AMI. The magnified views reveal finer structural differences and systematic alterations across the spectrum. These examples reflect recognizable ECG changes commonly associated with myocardial ischemia, injury, and infarction, including prominent T-wave changes, ST-segment elevation, evolving T-wave inversion, and pathological Q waves. These categories correspond to clinically recognized myocardial states, although individual recordings may not exhibit all classical features. The examples are illustrative across subjects and do not represent longitudinal changes within a single individual. Animated visualizations corresponding to each representative case are provided in Supplementary Movies 2–8, illustrating the synchronized evolution of ECG waveforms and the corresponding ECG “eyeball” throughout the 30-second recording.

Common-scale visualizations in Figure 4 demonstrate a consistent decrease in radial extent across conditions, while magnified views reveal finer structural distortions and progressive changes in morphology. These changes are consistent with recognizable ECG manifestations associated with myocardial ischemia, injury, and infarction (Figure 4), indicating that the “eyeball” representation captures clinically relevant electrophysiological alterations in a compact and visually interpretable form.

## Discussion

The present study introduces a novel ECG time–frequency “eyeball” framework that transforms a brief single-lead ECG recording into a visually interpretable geometric representation and a set of low-dimensional quantitative descriptors. The principal findings are threefold. First, the proposed “eyeball” metrics discriminated healthy subjects from patients with acute myocardial infarction (AMI), indicating that the ECG “eyeball” captures clinically meaningful information associated with AMI. Second, representative examples suggested a coherent morphologic continuum from normal patterns toward characteristic geometries consistent with myocardial ischemia, injury, and infarction, characterized by progressive contraction of the overall “eyeball,” altered compactness, and increasing contour irregularity (Figure 4). Third, the representation showed strong temporal robustness across short-term recordings (i.e., 30-second, 1-minute, 2-minute, and 5-minute). The “eyeball” metrics demonstrated minimal variation across recording durations in both cohorts, reflected by consistently high p-values using one-way ANOVA or Kruskal–Wallis test, as appropriate, and the morphology also remained visually preserved (see Supplementary Figures 1-3). Notably, the framework does not rely on explicit waveform delineation or fiducial point detection (e.g., P–QRS–T annotation), which may enhance robustness in short, noisy, or wearable-acquired ECG recordings. Importantly, the framework achieved these findings using only brief single-lead ECG recordings, suggesting the potential to capture clinically relevant electrophysiological alterations that may not be readily apparent during conventional ECG interpretation alone, while remaining compatible with wearable-style ECG acquisition. Taken together, these findings establish the ECG “eyeball” as an interpretable framework that transforms brief single-lead ECGs into stable geometric representations and low-dimensional quantitative descriptors, enabling visualization and quantification of clinically relevant dynamics.

The novelty of this work lies not simply in applying time–frequency analysis to ECG, but in converting that information into a disease-associated geometry that is simultaneously amenable to visual inspection and low-dimensional quantitative analysis. A key aspect of this framework is the use of the Hilbert transform to construct analytic signal trajectories from the intrinsic mode functions. The Hilbert transform does not alter the underlying oscillatory content of the intrinsic mode functions; rather, it converts each into an analytic signal whose complex-plane trajectory provides a geometric representation of its instantaneous dynamics. This enables subtle variations in cardiac electrical activity that may remain hidden during conventional ECG analysis to become both visually interpretable and quantitatively measurable, thereby facilitating the identification of hidden AMI signatures. Prior ECG time–frequency studies have shown the usefulness of wavelets, short-time Fourier approaches, and empirical mode decomposition for denoising, feature extraction, and automated classification^21^. Likewise, earlier geometric or phase-space approaches demonstrated that myocardial ischemia can distort ECG morphology in ways that are not fully captured by conventional ST-segment rules^11,23^. More recently, machine learning and deep learning studies have shown that ECGs can support detection of acute ischemia, myocardial injury, AMI, and structural heart disease with good diagnostic performance^6,7,11–13,15,18^. However, most such methods operate as high-dimensional feature engines or black-box predictors. In contrast, the current framework generates a recognizable morphology from a single lead and a short recording window, together with quantitative rotational and geometrical descriptors, providing a low-dimensional representation that complements both conventional ECG interpretation and AI-based analysis.

The observed geometric changes are physiologically plausible in the setting of myocardial ischemia and infarction. Acute myocardial ischemia alters resting membrane potential, action potential duration, conduction velocity, and the balance between depolarization and repolarization across the ischemic and non-ischemic myocardium^2–4^. These electrophysiological disturbances are often reflected on the ECG as hyperacute T-wave changes, ST-segment elevation or depression, evolving T-wave inversion, QRS distortion, and eventually the development of pathological Q waves and loss of viable electrical forces^1,3,4^. Because the proposed “eyeball” is built from data-adaptive oscillatory components rather than from fixed fiducial landmarks alone, it is reasonable to interpret it as a compact representation of the joint depolarization–repolarization behavior of the signal across multiple scales. In that sense, changes in “eyeball” morphology and its rotational features may reflect the dynamically evolving, integrated footprint of ischemic conduction delay, altered repolarization gradients, and infarct-related loss of organized myocardial activation.

One plausible biological explanation for the specific geometric evolution observed here is that progressive myocardial ischemia and infarction reduce the coherence and amplitude diversity of the underlying electrical signal while increasing regional heterogeneity. The reduction in overall “eyeball” size may reflect attenuation of organized excursions in the intrinsic mode function trajectories, consistent with diminished viable myocardium and redistribution of electrical forces. Increasing compactness may reflect collapse of the trajectories toward a narrower range of oscillatory states, whereas increasing irregularity may reflect spatially heterogeneous conduction slowing and dispersion of repolarization, which introduce asymmetry, local instability, and contour distortion into the “eyeball” representation (Figure 4). Importantly, this interpretation remains inferential. A definitive mechanistic mapping between individual “eyeball” features and specific electrophysiological processes will require dedicated experimental or multimodal validation with synchronized angiographic, biomarker, and imaging correlates^3,4^.

From a clinical perspective, the proposed framework is attractive for several reasons. Current acute coronary syndrome (ACS) pathways, particularly for acute myocardial infarction, still depend on rapid ECG acquisition, repeated interpretation, and integration with symptoms and biomarkers, yet subtle and dynamically evolving ischemic/occlusion-related patterns are easily overlooked, especially early or outside specialized settings^1,5,24^. A single-lead, short-recording representation that remains stable over 30 seconds could be useful as an adjunctive triage signal in emergency departments, ambulances, remote monitoring programs, and eventually wearable ecosystems. This is particularly relevant because contemporary smartwatches and handheld devices (or even emerging smart rings) already capture short single-lead ECGs (often around 30 seconds) and recent studies have shown growing diagnostic utility of such recordings for both structural and ischemic phenotypes^13,15,16,18,19,25^. At the same time, this framework should be viewed as a complement rather than a replacement for standard 12-lead ECG interpretation, high-sensitivity troponin algorithms, or guideline-directed ACS assessment. It could potentially enhance cardiovascular screening and early identification of patients with suspected AMI, enrich early risk stratification, and prompt referral for more definitive diagnostic testing.

Several strengths of the proposed framework deserve emphasis. The first is compatibility with brief single-lead ECG acquisition, which improves theoretical portability to prehospital and wearable scenarios. The second is temporal robustness; the duration analyses demonstrated minimal variation in the proposed “eyeball” metrics across short-term recordings (i.e., 30 seconds, 1 minute, 2 minutes, and 5 minutes), reflected by consistently high p-values (all p≥0.93-0.99, predominantly p>0.99; see Supplementary Figure 1 and Supplementary Figure 2). This indicates that extending recording duration produced little measurable change in the extracted features, supporting the practical sufficiency of brief recordings for this framework. In addition, the representative geometries remained qualitatively consistent across durations (Supplementary Figure 3). The third is visual interpretability. Unlike many AI-ECG approaches that primarily output a classification score, the “eyeball” framework affords an at-a-glance morphologic representation together with multiple quantitative descriptors that may be easier for clinicians and researchers to inspect, compare, and communicate. The fourth is that the framework provides both component-level and global descriptors of the ECG “eyeball”. Component metrics preserve detailed oscillatory information and demonstrated greater sensitivity to disease-associated changes, whereas global metrics integrate information across all intrinsic mode functions into compact energy-weighted descriptors. Together, these complementary metrics provide both detailed physiological characterization and compact quantitative summaries of ECG dynamics, offering flexibility for different analytical and clinical applications. The aforementioned strengths support the translational potential of the ECG “eyeball” framework for future wearable and point-of-care cardiovascular applications.

Several limitations should be acknowledged. First, the current study is not a prospective chest-pain triage study; therefore, its findings should not yet be interpreted as evidence of performance in the full real-world differential diagnosis of acute coronary syndromes. Second, the AMI cohort was recorded 24–48 hours after the index event, which primarily reflects the evolved acute phase rather than the earliest hyperacute minutes-to-hours phenotype and may therefore differ from very-early ischemic signatures. Third, the representative progression shown in Figure 4 is intended to illustrate a morphologic spectrum across clinically recognized myocardial conditions and should not be interpreted as a formally validated multi-class classification framework. Fourth, translation to consumer wearable devices should be made cautiously, as performance may vary with signal quality, physiological variability, and real-world confounders.

Future work should therefore proceed along several parallel tracks. The highest priority is external validation in independent multicenter datasets across acute care, ambulatory, and wearable settings, with rigorous subgroup analyses by age, sex, rhythm status, infarct territory, and device type. The second priority is prospective clinical validation in consecutive chest-pain cohorts, ideally with adjudicated endpoints that include AMI, occlusion MI, myocardial injury, and 30-day major adverse cardiovascular events. The third is methodological and clinical expansion, including fusion with conventional ECG intervals or troponin data, developing automated/AI-powered classification pipelines built on the combined “eyeball” features as an interpretable feature layer rather than a standalone visualization, and extension of the framework to other cardiac conditions such as cardiomyopathies, valvular heart disease, hypertensive heart disease, and heart failure phenotypes. The fourth is reporting and translational rigor. Future studies should prioritize rigorous study design, transparent reporting, and translational readiness, particularly as the framework is extended toward clinical decision-support and wearable applications^26–28^.

In summary, the ECG time–frequency “eyeball” framework condenses clinically relevant nonlinear ECG dynamics into a stable, interpretable, and low-dimensional geometric representation. The framework discriminated patients with AMI from healthy subjects, revealed hidden AMI signatures, and organized characteristic morphologies across a spectrum of myocardial conditions, while remaining robust for recordings as short as 30 seconds. This approach could provide a new bridge between classical ECG interpretation and modern data-driven cardiovascular diagnostics, laying the foundation for accessible single-lead cardiovascular screening using emerging smart wearables.

## Methods

### Clinical Databases

Data used for this study were obtained through an agreement with the Telemetric and Holter ECG Warehouse (THEW) of the University of Rochester, NY (http://thew-project.org/index.htm). We employed deidentified recordings from two ECG databases: the Healthy Subjects Database (THEW identification: E-HOL-03-0202-003) and the Acute Myocardial Infarction Subjects Database (THEW identification: E-HOL-03-0160-001). This study was determined by Salus IRB not to constitute regulated human-subjects research because only deidentified ECG data were used; therefore, formal IRB review was not required (Study ID 26447).

#### Healthy Subjects Database

The original study population consisted of 202 healthy subjects. Healthy individuals were eligible for enrollment if they had no overt cardiovascular disease or history of cardiovascular disorders (including stroke, transient ischemic attack, or peripheral vascular disease); no history of hypertension (>150/90 mmHg); no use of prescribed medications; and no chronic systemic illness such as diabetes mellitus, asthma, or chronic obstructive pulmonary disease. Subjects were not enrolled if they had been evaluated by a physician for cardiovascular-related symptoms, including chest pain, palpitations, or syncope, even if subsequently considered healthy. All participants had a normal physical examination and demonstrated sinus rhythm on a standard 12-lead ECG without suspicious abnormalities (e.g., signs of ventricular hypertrophy, T-wave inversion, or intraventricular conduction disturbances). In cases of suspicious ECG changes, a normal echocardiogram and normal exercise ECG testing were required. Pregnant individuals were excluded.

In this study, we excluded an additional 32 subjects due to age <18 years, significant noise or insufficient analyzable signal, lead misplacement, or the presence of premature atrial beats. Accordingly, 170 healthy subjects were included in the final analysis. Characteristics of the analyzed healthy population are summarized in Table 1.

#### Acute Myocardial Infarction Subjects Database

The original acute myocardial infarction (AMI) database consisted of 93 patients. AMI was diagnosed based on characteristic symptoms, along with supporting clinical and enzymatic markers, as defined in the original database. Symptoms included sudden chest pain (typically radiating to the left arm or left side of the neck), shortness of breath, nausea, vomiting, palpitations, sweating, and anxiety. Exclusion criteria included non-sinus rhythm (e.g., atrial flutter or fibrillation, pacemaker rhythm, atrioventricular block, or sick sinus syndrome); major comorbidities such as malignancy, severe hepatic, renal or cerebral disease; and prior coronary artery bypass graft (CABG) surgery. Patients with a history of non-CABG coronary revascularization (e.g., percutaneous transluminal coronary angioplasty, stent implantation, or atherectomy) were eligible.

In this study, only ECG recordings obtained 24–48 hours after the index myocardial infarction (ID-1) were included, as some patients also underwent a second recording predischarge (days 5–10; ID-2). We excluded an additional 13 subjects due to significant noise or insufficient analyzable signal, lead misplacement, arrhythmia, or missing ID-1 data. Accordingly, 80 AMI patients were included in the final analysis. Characteristics of the analyzed AMI cohort are summarized in Table 1.

In both databases, recordings were acquired using the SpaceLabs-Burdick digital Holter recorder (SpaceLab-Burdick, Inc., Deerfield, WI), with a three-lead pseudo-orthogonal configuration (x,y,z) and a sampling frequency of 200 Hz. Here, we used only Lead x (left-to-right axis), which approximates Lead I of the standard 12-lead ECG and is representative of single-lead configurations used in wearable and portable devices^15,17,18,29^ (e.g., Apple Watch, Kardia, or Cardio Ring). Short-term segments were extracted from continuous Holter recordings. Specifically, continuous 30-second segments were selected to mimic the duration of smartwatch or handheld ECG measurements (see “Data Preparation” section for more details).

### Formation of the ECG Time-Frequency “Eyeball”

Let *x*(*t*) denote an acquired single-lead ECG recording in time-series (e.g., Lead-I–equivalent axis via smart wearables, or handheld devices). The proposed “eyeball” framework maps temporal ECG dynamics into a compact rotational-morphological representation, enabling extraction of novel rotational and geometrical features while facilitating intuitive visualization and rapid interpretation. The framework proceeds as follows.

#### Decomposition of the ECG recording

The ECG recording is decomposed into a finite set of intrinsic mode functions (IMFs) using the empirical mode decomposition method:

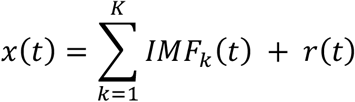

where *IMF*_*k*_(*t*) represents the *k*-th intrinsic mode function and *r*(*t*) is the residual component.

#### Derivation of Analytic Signals via Hilbert Transform

For each IMF, the analytic signal is obtained using the Hilbert transform^20,22^:

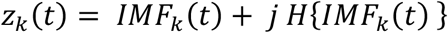

where *H*{·} denotes the Hilbert transform and 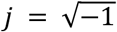.

#### Rotational Mapping in the Complex Plane

The temporal evolution of each analytic signal (***z_k_***(***t***)) defines a rotational trajectory in the complex plane, forming the basis of the proposed representation (Figure 1). This mapping is expressed as:

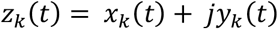

where *x*_*k*_(*t*) = *Re*(*z*_*k*_(*t*)) and *y*_*k*_(*t*) = *Im*(*z*_*k*_(*t*)) denote the real and imaginary components of each analytic signal.

The analytic signal further enables direct computation of instantaneous rotational features for each IMF trajectory, including instantaneous envelope (radial extent of the trajectory), instantaneous phase (angular position), and instantaneous frequency (angular velocity, or rotational speed). In polar form:

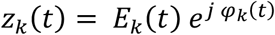

where *E*_*k*_(*t*) is the instantaneous envelope, and *φ*_*k*_(*t*) is the instantaneous phase. The instantaneous features are defined as:

- Instantaneous envelope (radial extent)

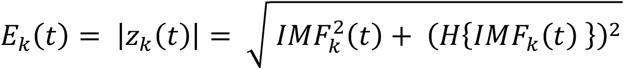
- Instantaneous phase (angular position)

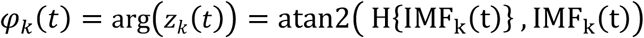

where atan2(.,.) denotes the two-argument arctangent function.
- Instantaneous frequency (angular velocity)

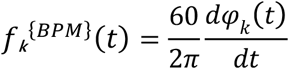

Instantaneous frequency is expressed in beats per minute (BPM) for clinical interpretability.

#### Construction of the ECG Time–Frequency “Eyeball”

Finally, the complex-plane trajectories from the first *N* analytic signals are superposed as:

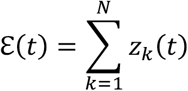

For *N*≥4, the resulting trajectory (ℰ(*t*)) forms a distinct eyeball-like geometry, termed the ECG Time–Frequency “Eyeball”. In the present study, *N*=4 was used for all analyses.

### Feature Extraction from the ECG Time–Frequency “Eyeball”

In total, twelve time-invariant features are extracted from the ECG “eyeball”, capturing its multi-component rotational dynamics, global rotational dynamics, and geometrical structure.

#### Component Rotational Features

Average instantaneous features across the ECG recording are computed for each IMF:

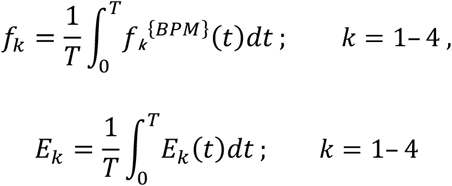

where *T* is the recording duration. These define the time-invariant, IMF-level rotational features for the “eyeball” framework:

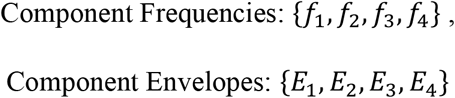

These averaged quantities can be interpreted as defining a circular approximation of each IMF trajectory, characterized by mean angular velocity and mean radial extent (dotted circles in Figure 1).

#### Global Rotational Features

To characterize the overall rotational dynamics of the ECG “eyeball” representation, we further defined two global instantaneous features (i.e., global instantaneous frequency and global instantaneous envelope) by combining information from the first four IMFs.

The global instantaneous frequency is defined as an energy-weighted average of the instantaneous frequencies of the first four IMFs:

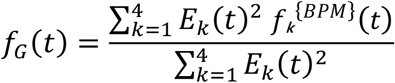

where *f*_*k*_^{*BPM*}^(*t*) and *E*_*k*_(*t*)^2^ denote the instantaneous frequency and instantaneous energy of the *k*-th IMF, respectively.

The global instantaneous envelope is defined as the root-sum-square envelope of the first four IMFs, thereby preserving the aggregate instantaneous energy within the combined “eyeball” representation:

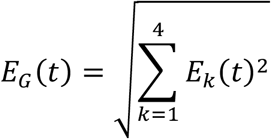

These global instantaneous features (shown in Figure 1) are then averaged across the ECG recording to extract time-invariant, global rotational features of the ECG “eyeball”:

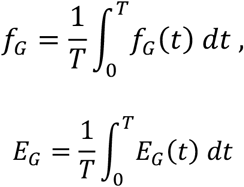

Such global descriptors summarize the overall rotational behavior of the ECG “eyeball” and complement the component rotational features. Global rotational features are summarized as:

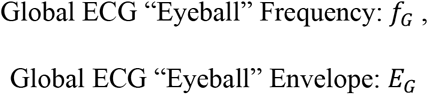

#### Geometrical Features

For geometric analysis, the “eyeball” morphology should be first reinforced using arc-length-based interpolation to ensure uniformly spaced points (one point per unit arc length) and avoid bias due to non-uniform point density (e.g., sparse points in high-curvature regions such as QRS-related components; Figure 1).

To quantify the global morphological distribution, centroid locations along the real (x-axis) and imaginary (y-axis) dimensions of the “eyeball” geometry are computed as:

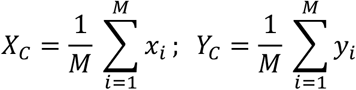

where (*x*_*i*_, *y*_*i*_) are points on the reinforced “eyeball” geometry, and *M* is the total number of points.

These centroid coordinates define the geometrical features of the “eyeball” framework as:

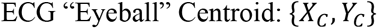

### Data Preparation

As the database provides long-term ambulatory recordings, we extracted 30-second ECG epochs during stable resting periods, targeting low-activity or sleep-like states. These states are minimally affected by motion artifacts and external stimuli, making them ideal for our morphology-based ECG analysis. To ensure consistency across all subjects, we: (1) identified the 6-hour interval with the lowest average heart rate (putative nocturnal rest) using a moving average over RR intervals, shifted in 15-minute steps; (2) selected the beginning of hour 2 within this 6-hour interval; and (3) extracted a stable 30-second epoch from that time. All 30-second ECG epochs were preprocessed using median filtering, whereby a 1-second median filter was applied and subtracted from the signal to remove baseline drift.

### Statistical Analysis

Continuous variables are presented as mean ± standard deviation (SD) and as median with interquartile range (IQR), unless otherwise stated. Categorical variables are summarized as counts and percentages. Differences between healthy and AMI cohorts for each metric were assessed using an unpaired t-test or the Mann–Whitney U test, as appropriate. A two-sided p-value < 0.05 was considered statistically significant.

The association between each ECG “eyeball” feature and the odds of AMI was evaluated using logistic regression models. For each feature, three models were constructed: an unadjusted model, an age- and sex-adjusted model, and a fully adjusted model including age, sex, and heart rate (using average NN interval). To facilitate comparisons across features, results are reported as normalized odds ratios (i.e., the odds ratio per one standard deviation increase in each feature), along with 95% confidence intervals (95% CI). Confidence intervals were computed using the profile likelihood method. To assess the incremental contribution of each feature beyond clinical covariates, likelihood-ratio tests were performed by comparing a reduced model (including age and sex, with or without heart rate) to a full model that additionally included the eyeball feature.

The discriminative ability of each feature for identifying AMI was evaluated using receiver operating characteristic (ROC) analysis. The area under the ROC curve (AUC) was computed as the performance metric. All statistical analyses were performed using Python 3.11.2 and R version 4.5.2.

## Data Availability

All data produced in the present study are available upon reasonable request to the authors.

## Acknowledgements

Rashid Alavi acknowledges support from the James Boswell postdoctoral fellowship at the California Institute of Technology (Caltech).

